# Investigating the relationship between sleep disturbances and white matter hyperintensities in older adults on the Alzheimer’s disease spectrum

**DOI:** 10.1101/2023.04.13.23288544

**Authors:** Farooq Kamal, Cassandra Morrison, Mahsa Dadar

**Affiliations:** Department of Psychiatry, McGill University, Montreal, Quebec, H3A 1A1, Canada; Douglas Mental Health University Institute, Montreal, Quebec, H4H 1R3, Canada; Department of Neurology and Neurosurgery, Faculty of Medicine, McGill University, Montreal, Quebec, H3A 2B4, Canada; McConnell Brain Imaging Centre, Montreal Neurological Institute, McGill University, Montreal, Quebec, H3A 2B4, Canada

**Keywords:** Older adults, Mild cognitive impairment, Alzheimer’s disease, Cognitive decline, White Matter Hyperintensities, Sleep disturbances, Amyloid positivity

## Abstract

**Background:** While studies report that sleep disturbance can have negative effects on brain vasculature, its impact on cerebrovascular disease such as white matter hyperintensities (WMHs) in beta-amyloid positive older adults remains unexplored.

**Methods:** Linear regressions, mixed effects models, and mediation analysis examined the cross-sectional and longitudinal associations between sleep disturbance, cognition, and WMH burden, and cognition in normal controls (NCs), mild cognitive impairment (MCI), and Alzheimer’s disease (AD) at baseline and longitudinally.

**Results:** People with AD reported more sleep disturbance than NC and MCI. AD with sleep disturbance had more WMHs than AD without sleep disturbances. Mediation analysis revealed an effect of regional WMH burden on the relationship between sleep disturbance and future cognition.

**Conclusion:** These results suggest that WMH burden and sleep disturbance increases from aging to AD. Sleep disturbance decreases cognition through increases in WMH burden. Improved sleep could mitigate the impact of WMH accumulation and cognitive decline.

## 1 Introduction

Sleep disturbance refers to experiencing difficulties when falling and/or staying asleep throughout the night. Such disturbances may result in daytime fatigue and may also cause other related problems, such as concentration issues, memory loss, irritability, and depression.^1^ Sleep disturbances such as insomnia or sleep apnea are a physiological hallmark of increasing age and become more prevalent in people with mild cognitive impairment (MCI) and Alzheimer’s disease (AD).^2–5^ Sleep disorders observed in people with AD^6^ and MCI^7^ are more severe than healthy older adults. For example, people with AD and MCI tend to experience more disrupted nocturnal sleep, earlier bedtimes, more variable wake up times, and fragmented sleep, compared to healthy older adults.^6–8^ Sleep disturbances in aging, MCI, and AD have been observed to be related to increased cognitive change, pathology, and risk/progression of AD.^2^

With respect to cognition, several studies have noted a positive correlation between the severity of sleep disturbance and the severity of cognitive decline in healthy older adults ^9^ and individuals with MCI/AD.^4,5,10,11^ These declines are often observed in global cognition and memory.^10,11^ Additionally, sleep disturbances have been thought to be associated with the accumulation of amyloid-beta (Aβ)^4,5^ and neurodegeneration.^12^ Higher levels of Aβ deposition have been observed in people with more sleep disruptions.^4,13–15^ The presence of Aβ is observed in people with AD, and is used as a biomarker to identify individuals who will develop AD.^16^ Therefore, the relationship between Aβ and sleep highlights the crucial role that sleep plays in the development of AD-related pathology. Other pathological changes in the brain, such as white matter hyperintensities (WMHs), have also been associated with sleep disturbances. For example, insomnia and short sleep durations are associated with increased WMH burden.^17–19^ However, the results examining this relationship have not been consistently reported, with some studies finding no association between sleep disturbance and WMH burden.^20,21^ WMHs are observed as increased signal during T2-weighted or fluid-attenuated inversion recovery (FLAIR) magnetic resonance imaging (MRI) scans. They are often used as a proxy for cerebrovascular disease, which is a well-established contributor to cognitive decline and dementia.^22–24^ WMHs are associated with increased cognitive decline in healthy older adults^25–27^ and in MCI^26,28–30^ as well as increase risk of conversion to MCI and dementia.^31^

The current research investigating the relationship between sleep and WMHs in aging and dementia has several limitations that may account for conflicting results. Previous research has not yet examined regional WMH differences and sleep disturbance in individuals who are amyloid positive. Healthy older adults and individuals with MCI who are amyloid positive are on the AD trajectory, therefore this difference in diagnostic outcome may influence the relationship between sleep and WMH burden. Most studies have used a total brain WMH approach, which may not capture regional differences specific to sleep disturbances. It is possible that sleep may be associated with WMH accumulation in specific regions, therefore a regional approach would provide insight into the relationship between sleep and regional WMHs. Other findings have reported that posterior WMHs are associated with AD^32,33^ while a widespread accumulation of WMH is associated with vascular dementia.^34^ Thus, a regional approach may indicate whether sleep disturbances are more strongly associated with AD-related WMH burden or vascular-related WMH burden.

The goal of this study was to examine whether sleep disturbances influence WMH burden in healthy older adults, people with MCI, and AD who were amyloid positive, and whether these sleep disturbances influence global cognition. To our knowledge, this would be the first study to examine the relationship between sleep disturbances and WMH burden and global cognition in older adults who are amyloid positive. Given that sleep disturbances may be modifiable or treatable in older adults using various intervention techniques (e.g., medications, cognitive-behavioral strategies, and circadian therapies),^35,36^ reducing cognitive decline and pathological brain changes may be mitigated by treating sleep disturbance.

## 2 Methods

### 2.1 Alzheimer’s Disease Neuroimaging Initiative

The data used for this article were taken from the ADNI database (adni.loni.usc.edu), which was established in 2003 as a public-private venture with Michael W. Weiner, MD as the Principal Investigator. ADNI’s primary aim is to determine if serial magnetic resonance imaging (MRI), positron emission tomography (PET), other biological markers, and clinical and neuropsychological assessment can be utilized to measure the progression of MCI and AD. The research was granted ethical approval from the review boards of all the involved institutions, and written consent was obtained from participants or their study partner. The participants for this study were taken from all the ADNI cohorts (ADNI-1, ADNI-2, ADNI-GO, and ADNI-3).

### 2.2 Participants

Participant inclusion and exclusion criteria are available at www.adni-info.org. All Participants were included in the study if they were between the ages of 55 and 95 and did not display any signs of depression. Healthy older adults did not exhibit memory decline or impaired global cognition, as assessed by the Wechsler Memory Scale, Mini-Mental Status Examination (MMSE) and Clinical Dementia Rating (CDR). MCI participants had MMSE scores between 24 and 30, CDR scores of 0.5, and abnormal scores on the Wechsler Memory Scale. AD participants had abnormal memory function on the Wechsler Memory Scale, MMSE scores between 20 and 26, a CDR of 0.5 or 1.0, and met the National Institute of Neurological and Communicative Disorders and Stroke and the Alzheimer’s Disease and Related Disorders Association criteria.^37^

To make sure that the participants in this study were on the AD trajectory, participants were only included if they had evidence of amyloid positivity. PET and CSF values were both used to make this determination. Amyloid positivity was identified based on the following criteria: 1) a standardized uptake value ratio (SUVR) of > 1.11 on AV45 PET^38^, 2) an SUVR of >1.2 using Pittsburgh compound-B PET,^39^ 3) an SUVR of ≥1.08 for Florbetaben (FBB) PET,^40^ or 4) a cerebrospinal fluid Aß1-42 ≤ 980 pg/ml as per ADNI recommendations.

Participant sleep scores were also downloaded from the ADNI website. Consistent with previous research, the presence of sleep disturbance was determined by caregiver report on the Neuropsychiatric Inventory (NPI), a brief questionnaire form of the NPI (NPI-Q), or Diagnosis and Symptoms Checklist (ADSXLIST).^41–43^ The NPI is a caregiver-informant interview that assess behavioral changes including sleep problems. The NPI-sleep questionnaire pertains to recent changes in the patient’s sleep behavior. The initial question determines the presence or absence of abnormal sleep changes. In the present study, we focused on questions (from all three questionnaires) that were specifically related to sleep and excluded questions that assessed erratic behavior. That is, we kept questions relating to: difficulties in falling asleep (K1), getting up during the night (K2), awakening the caregiver during night (K4), waking up too early in the morning (K6), and excessive daytime napping (K7). In the ADSXLIST, we extracted information on patient insomnia. As for the NPI, three questions were included asking a caregiver if the patient: awakens during night, arises too early in the morning, or takes excessive naps during the day. These questions were scored as either zero for the absence and one for the presence of each specific symptom. This approach ensured that we were able to accurately identify and include participants who were experiencing sleep disturbance, while minimizing the inclusion of individuals who may have been experiencing other types of disruption.

A total of 912 participants with amyloid positivity were included in this study [198 healthy controls (NC), 504 MCI, and 210 AD]. Out of the 912 participants, 292 had sleep disturbances (47 NC, 159 MCI, and 86 AD). The groups had the following number of average follow-up time: 1) NC: 4.00 years for no sleep disturbance and 4.49 years with sleep disturbance, 2) MCI: 3.18 years for no sleep disturbance and 3.14 years with sleep disturbance, 3) AD: 1.42 years for no sleep disturbance and 1.45 years with sleep disturbance. Only participants that had at least two follow-up visits were included in the study. To ensure that follow-up time duration did not impact our study findings, the models were repeated with the groups matched based on sleep disturbance and follow-up time (i.e., excluding additional follow-up timepoints from groups that had longer follow-up durations).

### 2.3 Structural MRI acquisition and processing

All participants underwent MRI scanning using standardized acquisition protocols designed and implemented by ADNI. Further details about the MRI protocols and imaging parameters can be found at http://adni.loni.usc.edu/methods/mri-tool/mri-analysis/. MRI data from all participants (baseline and longitudinal) was downloaded from the ADNI public website.

All T1w scans were pre-processed using our standard pipeline which includes: noise reduction, ^44^ intensity inhomogeneity correction, ^45^ and intensity normalization into range [0-100]. The pre-processed images were then linearly (9 parameters: 3 translation, 3 rotation, and 3 scaling) (Dadar et al., 2018) registered to the MNI-ICBM152-2009c average template.^47^

### 2.4 WMH measurements

A well-established, validated method for segmenting WMHs in aging and neurodegenerative diseases was used to obtain WMH measurements.^48^ This technique has been used in multi-center studies^49,50^ including the ADNI cohort.^31^ WMHs were automatically segmented using the T1w contrast, in addition to location and intensity features from a library of manually segmented scans (50 ADNI participants that were not part of the study) in combination with a random forest classifier to detect the WMHs in new images.^48^ WMHs were segmented using T1w images (instead of FLAIR and T2w/PD scans) because ADNI1 only acquired T2w/PD images with resolutions of 1×1×3 mm^3^, whereas ADNI2/GO acquired only axial FLAIR images with resolutions of 1×1×5 mm^3^, and ADNI3 acquired sagittal FLAIR images with resolutions of 1×1×1.2 mm^3^. Due to the potential inconsistencies in WMH measurements across the ADNI cohorts, we opted to include consistently acquired T1w images to measure WMH burden. These T1w-based WMH volumes have been previously observed to be very highly correlated with FLAIR and T2w based WMH loads in the ADNI dataset.^46,51^ A visual assessment (performed by M.D.) was conducted to assess the quality of the registrations and WMH segmentations. Cases that did not meet the quality control criteria were removed from the analyses (N = 59). The WMH load was determined as the volume of all voxels identified as WMH in the standard space (in mm^3^) and was therefore normalized for head size. Regional and total WMH volumes were calculated based on Hammers Atlas.^51,52^ Analyses were completed separately for frontal, temporal, parietal, occipital, total WMHs. Regional WMH values (i.e., frontal, temporal, parietal, and occipital) were summed across the right and left hemispheres to obtain one score for each region. To achieve normal distribution, All WMH volumes were log-transformed.

### 2.5 Global Cognition

The Clinical Dementia Rating – Sum of Boxes (CDR-SB) was included as a measure of global cognitive functioning. This cognitive score was also downloaded from the ADNI website.

### 2.6 Statistical Analysis

#### 2.6.1 Baseline Assessments

Participant demographic information is presented in Table 1. T-tests and chi-square analyses were performed on the demographic information and corrected for multiple comparisons using Bonferroni correction.

**Table 1:** Demographic and clinical characteristics for NCs, MCI and AD

| Demographic Information | NC<br>n = 198 | MCI<br>n = 504 | AD<br>n = 210 |
| --- | --- | --- | --- |
| Baseline Age | 75.0 ± 5.2 | 73.1 ± 7.1 | 74.4 ± 7.8 |
| Education | 16.1 ± 2.6 | 16.0 ± 2.9<sup>1</sup> | 15.6 ± 2.7<sup>1</sup> |
| APOE ε4+ | 78 (39%) | 313 (62%)<sup>1</sup> | 158 (75%)<sup>1,2</sup> |
| Sleep Disturbance | 47 (24%) | 159 (31%)<sup>1</sup> | 86 (41%)<sup>1,2</sup> |
| Male sex | 91 (46%) | 298 (59%)<sup>1</sup> | 117 (55%)<sup>1</sup> |
| BMI | 26.5 ± 4.5 | 26.7 ± 4.7 | 25.3 ± 4.1<sup>1</sup> |
| Hypertension | 96 (48%) | 238 (47%) | 103 (49%) |
| Diabetes | 13 (7%) | 36 (7%) | 15 (7%) |
| Baseline CDRSB | .04 ± .2 | 1.55 ± .9<sup>1</sup> | 4.44 ± 1.6<sup>1,2</sup> |

*Notes:* values are expressed as mean ± standard deviation, or number of participants (percentage %). Statistically significant results reported if they survived Bonferroni correction. <sup>1</sup> = statistically significant difference from NC. <sup>2</sup> = statistically significant difference from MCI. BMI = Body Mass Index. APOE $\epsilon$ 4+ = number and percentage of people with at least one $\epsilon$ 4 allele. NC = normal control. MCI = mild cognitive impairment. AD = Alzheimer's disease.

The following linear regression models were used to investigate the impact of sleep differences on total, regional WMH load (frontal, temporal, parietal, occipital, and total) within each diagnostic group at baseline. The models included age, years of education, sex, and APOE4 status, as covariates. The categorical variable of interest was Sleep disturbance (i.e., 0 = no, 1 = yes).

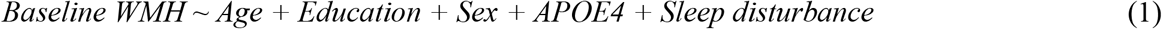

#### 2.6.3 Longitudinal Assessments

Longitudinal linear mixed-effects models were used to investigate whether there is an interaction between diagnostic group and sleep status, impacting regional WMH loads (frontal, temporal, parietal, occipital, and total). The models included age, years of education, sex, and APOE4 status as covariates. The categorical variables of interest were Sleep disturbance and its interaction with Diagnosis, based on baseline diagnosis. Participant ID was included as a categorical random effect.

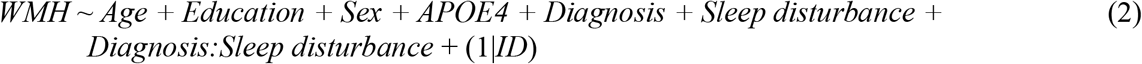

The following linear mixed effects model was conducted to examine whether sleep disturbance would influence global cognition (i.e., CDR-SB). The models included age, years of education, sex, and APOE4 status as covariates. The categorical variable of interest was Sleep disturbance.

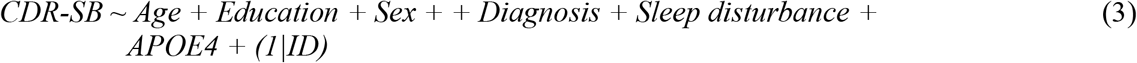

Finally, we used mediation analysis to test the hypothesis that WMH burden mediates the impact of sleep disturbance on future cognitive decline, including age, sex, years of education, and APOE4 status as covariates. Future cognitive decline was measured as yearly rate of change in CDR-SB scores in participants that had at least 3 longitudinal timepoints. The mediation analysis was performed using the mediation toolbox by Wager et al. (2009) (https://github.com/canlab/MediationToolbox). A 95% bootstrap confidence interval based on 10,000 bootstrap samples was used to estimate significance.

All continuous values were z-scored within the population prior to the regression and mediation analyses. All results were corrected for multiple comparisons using false discovery rate (FDR), p-values are reported as raw values with significance then determined by FDR correction.^53^ All statistical analyses were performed using MATLAB version 2021a. To complete the baseline analysis, we used function fitlm and for longitudinal assessments we used fitlme.

## 3 Results

### 3.1 Demographics and Cognitive Scores

Table 1 presents demographic and clinical information of participants in the study. Differences in demographics are reported for those that survived correction for multiple comparisons. The AD group had more people with sleep disturbance (41%) than both MCI (31%) and NC (24%), and the MCI group had significantly more people with sleep disturbance than NC. NCs had significantly greater education levels than AD *(t*=3.17, *p*=.001). NCs had fewer people with APOE □4+ than both MCI (x^2^= 28.79, *p*<.001) and AD (x^2^=52.24, *p*<.001). MCI had more people with APOE □ 4+ than AD (x^2^= 10.81, *p*=.001). NC had significantly greater body mass index (BMI) than AD (x^2^=3.36, *p*<.001). MCI also had g BMI than AD (x^2^=3.50, *p*<.001). There were no differences in the vascular risk factors of hypertension or diabetes between the groups. Baseline cognitive performance decreased with each stage of progressive decline for the CDRSB (NC:MCI, *t=-*35.41, *p*<.001; NC:AD, *t=-*39.75, *p*<.001; MCI:AD, *t* = -24.70, *p*<.001).

### 3.2 Baseline Assessments

Figure 1 shows boxplots of baseline WMHs overall and separately for each lobe. As expected, AD groups had significantly greater WMH burden than the NC group overall and across all regions (*t* belongs to [4.56–3.06], *p*<.01, Figure 1). MCI had significantly greater WMH burden than the NC group only in the frontal region (*t* = 2.15, *p*=.03). People with AD had significantly greater WMH burden than the MCI group in all regions (*t* belongs to [3.46–2.21], *p*<.03). In addition, people with AD who reported sleep disturbance had significantly greater WMH loads at total, frontal, and occipital regions than people with AD without sleep disturbance (*t* belongs to [2.11–3.12], *p*<.03). No significant differences were observed in MCI and NC groups.

**Figure 1.**
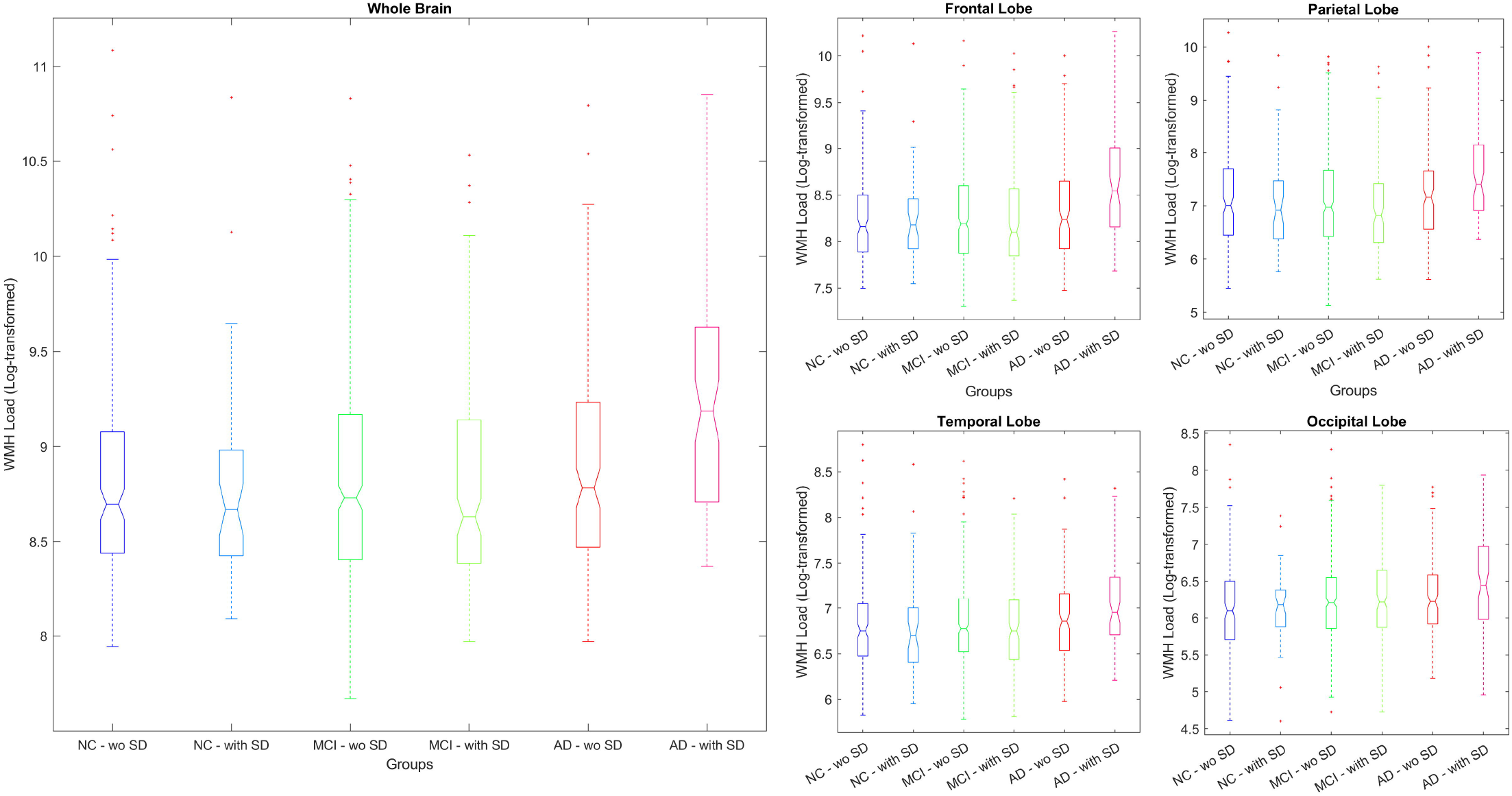
Boxplots showing baseline WMH distributions across diagnostic groups for each lobe. NC = normal controls. MCI = Mild cognitive impairment. AD = Alzheimer’s Disease. WMH = white matter hyperintensity. SD = Sleep Disturbance. wo = without.

### 3.3 Longitudinal Assessments

Table 2 summarizes the results of the longitudinal linear mixed effects models, revealing significant interactions between a diagnosis of AD and sleep disturbance. Specifically, people with AD who reported sleep disturbance had significantly increased WMH loads in all regions except occipital compared to both NC (*t* belongs to [2.58–2.44], *p*<.02) and people with MCI (*t* belongs to [3.18–2.28], *p*<.03).

**Table 2:** Linear mixed model results showing the interaction between diagnostic group (NC, MCI, and AD) and sleep disturbance.

|  | Total |  | Frontal |  | Temporal |  | Parietal |  | Occipital |  |
| --- | --- | --- | --- | --- | --- | --- | --- | --- | --- | --- |
|  | *T* stat | *P* | *T* stat | *P* | *T* stat | *P* | *T* stat | *P* | *T* stat | *P* |
| MCI vs NC:<br>Sleep Disturbance | .46 | =.654 | .12 | =.902 | .368 | .071 | .75 | .450 | -.11 | .910 |
| AD vs NC: Sleep<br>Disturbance | 2.74 | **=.006\*** | 2.58 | **=.009\*** | 2.44 | **=.015\*** | 2.44 | **=.01\*** | .91 | .363 |
| AD vs MCI:<br>Sleep Disturbance | 2.99 | **=.003\*** | 3.18 | **=.001\*** | 2.71 | **=.006\*** | 2.28 | **=.02\*** | 1.29 | .195 |

*Notes:* \* Represents results that are significant when uncorrected and bolded values represent those that remained significant after correction for multiple comparisons.

Additional exploratory analyses were completed within the AD group, to investigate whether the longitudinal mixed effects model results (Model 2) remained consistent without contrasting AD patients against NC and MCI. The within-group models also showed significantly greater WMH loads in AD patients with sleep disturbance than AD patients without sleep disturbance in total, frontal, and occipital lobes (*t* belongs to [2.56–3.99], *p*<.01).

#### 3.3.1 Global Cognition and regional WMH

The longitudinal models showed a significant impact of sleep disturbance on global cognition, as measured by CDR-SB (*t* = 2.71, *p* = .007), suggesting that sleep disturbance (compared to no sleep disturbance) is associated with increased CDRSB scores (i.e., worse cognition). Table 3 summarizes the results of the mediation analyses investigating whether the relationship between sleep disturbance and future cognitive decline is mediated by WMH burden. The results supported the existence of a mediation effect of WMH load on the relationship between sleep disturbance and future cognitive decline for total, frontal, and occipital WMHs, suggesting that sleep disturbance increases cognitive deficits through increases in WMH burden.

**Table 3:**
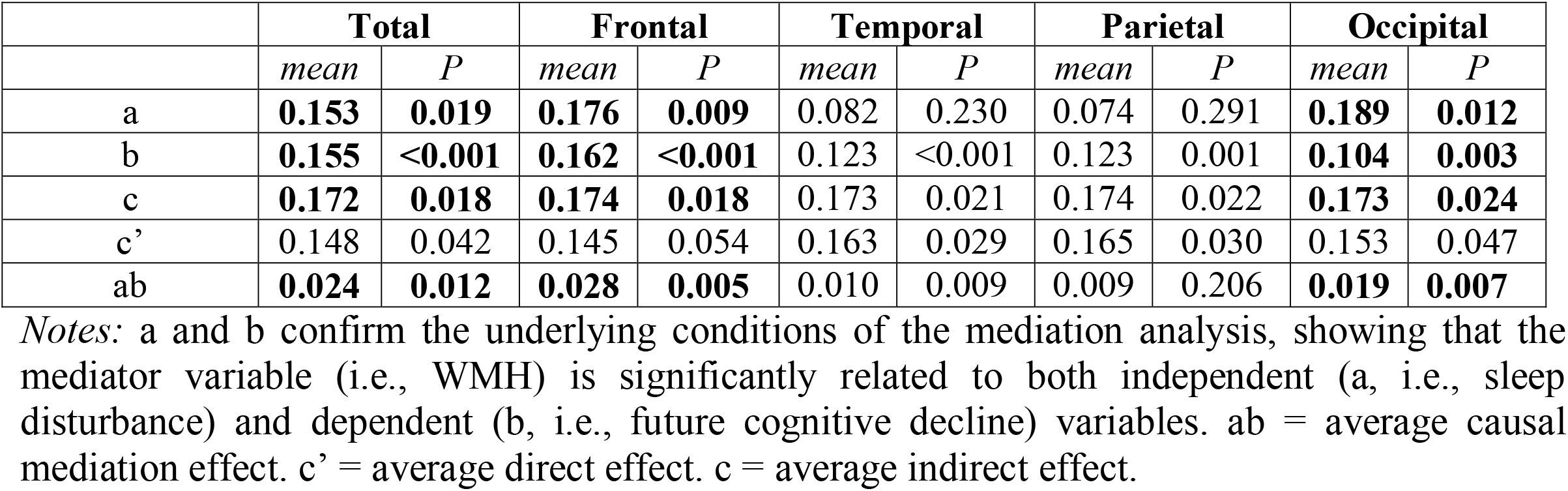
Mediation analysis results showing a mediation effect of WMH burden on the relationship between sleep disturbance and future cognition.

|  | **Total** |  | **Frontal** |  | **Temporal** |  | **Parietal** |  | **Occipital** |  |
| --- | --- | --- | --- | --- | --- | --- | --- | --- | --- | --- |
|  | *mean* | *P* | *mean* | *P* | *mean* | *P* | *mean* | *P* | *mean* | *P* |
| a | **0.153** | **0.019** | **0.176** | **0.009** | 0.082 | 0.230 | 0.074 | 0.291 | **0.189** | **0.012** |
| b | **0.155** | **<0.001** | **0.162** | **<0.001** | 0.123 | **<0.001** | 0.123 | 0.001 | **0.104** | **0.003** |
| c | **0.172** | **0.018** | **0.174** | **0.018** | 0.173 | 0.021 | 0.174 | 0.022 | **0.173** | **0.024** |
| c' | 0.148 | 0.042 | 0.145 | 0.054 | 0.163 | 0.029 | 0.165 | 0.030 | 0.153 | 0.047 |
| ab | **0.024** | **0.012** | **0.028** | **0.005** | 0.010 | 0.009 | 0.009 | 0.206 | **0.019** | **0.007** |

*Notes:* a and b confirm the underlying conditions of the mediation analysis, showing that the mediator variable (i.e., WMH) is significantly related to both independent (a, i.e., sleep disturbance) and dependent (b, i.e., future cognitive decline) variables. ab = average causal mediation effect. c' = average direct effect. c = average indirect effect.

## Discussion

Sleep disturbances are known to increase cognitive decline, AD-related pathology development and risk for AD.^2^ However, the relationship between sleep disturbances and WMHs, a known risk factor of cognitive decline and AD, is relatively unexplored. The current study investigated this association between sleep disturbances and WMHs in amyloid-positive older adults. Furthermore, the impact of sleep disturbances on cognitive functioning was also examined. At baseline, we observed that AD patients with sleep disturbance had significantly larger WMH burden than those without sleep disturbance. Longitudinal results revealed differences between people with AD with sleep disturbance compared to NCs with no sleep disturbance at all regions, except occipital. Furthermore, longitudinal analysis revealed that people with AD with sleep disturbance had increased WMH than MCI. Sleep disturbance was also related to global cognition (as measured by the CDR-SB). Finally, we found a mediation effect of total, frontal, and occipital WMH burden on the relationship between sleep disturbance and future cognition. Taken together, these findings suggest that sleep disturbances are positively associated with WMH accumulation in older adults on the AD trajectory and lead to cognitive deficits through increase in WMH burden.

The results of the current study are consistent with prior research demonstrating a link between sleep disturbance and WMH. Several previous studies with older adults have reported similar results, supporting the notion that sleep disturbances play an important role in WMH burden.^17–19^ It should be noted that some studies report no association between sleep disturbance and WMH burden.^20,21^ Li et al. (2020) did not specifically investigate the relationship between sleep and WMH but rather focused on the mediating effect of WMH on the relationship between sleep and depression. Therefore, the lack of significant association between sleep and WMH in this study may be due to the use of a sample of older adults with depression, who may have different underlying brain changes than older adults without depression. Additionally, the study did not examine longitudinal data but instead looked at cross-sectional data only. Similarly, Zitser et al. (2020), did not find a significant association between sleep duration and WMH in a large cohort of middle-aged and older individuals over a period of 28 years. However, the study did not examine regional WMH but rather looked at global WMH burden, which may have masked potential regional differences. The authors also noted that their sample had a relatively small proportion of individuals with poor sleep, which may have limited the statistical power to detect significant associations.^21^ Thus, the present study utilizes longitudinal data to provide precise estimates of the association between Aβ pathology and self-reported sleep duration on WMH burden. This study builds upon smaller baseline studies that have observed this association inconsistently to date.

The results of this study are also consistent with previous reports which found more severe sleep disturbances in people with AD^6^ and MCI^7^ compared to healthy older adults. However, we did not observe a diagnosis by sleep disturbance interaction for MCI (vs healthy older adults), indicating that the relationship between sleep disturbance and WMH burden is not significantly different in people with MCI compared to healthy older adults. On the other hand, we did observe that people with AD with sleep disturbance had increased WMH burden at baseline compared to healthy older adults with and without sleep disturbances. Over time, we also observed that people with AD had increased WMH compared to both healthy older adults and people with MCI with and without sleep disturbances. These findings follow previous reports indicating that WMH burden is more strongly associated with sleep disturbances in AD.

Sleep disturbance was also observed to be associated with changes in global cognition functioning (as measured by the CDR-SB). This relationship is consistent with previous reports indicating that disturbances in sleep are negatively associated with cognitive functioning.^10,11^ These findings indicate that sleep disturbances are associated with future cognitive decline in older adults on the AD trajectory. Thus, it may be possible to reduce cognitive decline in older adults by managing sleep disturbances. However, it is important to note that future research is needed to examine whether these sleep changes contribute to AD progression or simply reflect early symptoms. In addition, it is possible that different sleep disturbances (e.g., including sleep fragmentation, abnormal sleep duration, insomnia, excessive daytime sleepiness, sleep-disordered breathing), are associated with varying risks of pathological changes and may influence AD in different manners. Previous conflicting findings regarding the association of sleep with WMHs could be attributed to an inconsistent definition of sleep disturbance as well as subjective reports of sleep disturbance. For instance, research examining sleep disturbance typically incorporates a wide range of day and night-time abnormalities from self-reported sleep characteristics. This limitation could lead to the inclusion of individuals who do not actually experience sleep disturbance in the sleep disturbance group, which would contribute to the conflicting findings reported in the literature.

In conclusion, sleep disturbances increase in prevalence in individuals who experience cognitive decline, particularly in AD. These sleep disturbances are associated with decreased quality of life for both patients and caregivers. There is also growing evidence that sleep disturbances may be linked to Aβ accumulation and increased atrophy. The current study added to this literature by observing that sleep disturbances are associated with WMH burden in older adults on the AD trajectory. This finding suggests that individuals on the AD trajectory, who experience faster WMH progression,^30^ may also develop a more widespread accumulation of WMH if they experience sleep disturbances. Therefore, if sleep disturbances are managed, it may be possible to reduce the WMH burden observed in people with AD (in particular for frontal WMHs which have a more vascular origin),^54^ thus reducing cognitive change and slowing disease progression. Identifying and treating sleep disturbances in individuals with dementia may have important implications for disease management and may help develop new interventions to help mitigate disease progression.

## Data Availability

All data produced are available online at https://adni.loni.usc.edu/

## Acknowledgments

Data collection and sharing for this project was funded by the Alzheimer’s Disease Neuroimaging Initiative (ADNI) (National Institutes of Health Grant U01 AG024904) and DOD ADNI (Department of Defense award number W81XWH-12-2-0012). ADNI is funded by the National Institute on Aging, the National Institute of Biomedical Imaging and Bioengineering, and through generous contributions from the following: AbbVie, Alzheimer’s Association; Alzheimer’s Drug Discovery Foundation; Araclon Biotech; BioClinica, Inc.; Biogen; Bristol-Myers Squibb Company; CereSpir, Inc.; Cogstate; Eisai Inc.; Elan Pharmaceuticals, Inc.; Eli Lilly and Company; EuroImmun; F. Hoffmann-La Roche Ltd and its affiliated company Genentech, Inc.; Fujirebio; GE Healthcare; IXICO Ltd.; Janssen Alzheimer Immunotherapy Research & Development, LLC.; Johnson & Johnson Pharmaceutical Research & Development LLC.; Lumosity; Lundbeck; Merck & Co., Inc.; Meso Scale Diagnostics, LLC.; NeuroRx Research; Neurotrack Technologies; Novartis Pharmaceuticals Corporation; Pfizer Inc.; Piramal Imaging; Servier; Takeda Pharmaceutical Company; and Transition Therapeutics. The Canadian Institutes of Health Research is providing funds to support ADNI clinical sites in Canada. Private sector contributions are facilitated by the Foundation for the National Institutes of Health (www.fnih.org). The grantee organization is the Northern California Institute for Research and Education, and the study is coordinated by the Alzheimer’s Therapeutic Research Institute at the University of Southern California. ADNI data are disseminated by the Laboratory for Neuro Imaging at the University of Southern California.

